# Prevalence and factors associated with post-partum depression in a remote area of Cameroon: a cross-sectional study

**DOI:** 10.1101/2022.04.12.22273774

**Authors:** Therence Nwana Dingana, Stewart Ndutard Ngasa, Neh Chang Ngasa, Leticia Armelle Sani Tchouda, Christabel Abanda, Juste Niba, Bill Erich Mbanyor, Eric Wah Sanji, Carlson-Sama Babila

## Abstract

Postpartum depression is one of the many challenges associated with childbirth. In Cameroon, the focus is more on postpartum obstetric complications resulting in underdiagnosis and misdiagnosis of this condition. The current socio-political crisis plaguing the English-speaking part of Cameroon has increased the stressors that may inherently increase the prevalence. There is no published data describing post-partum depression in a rural setting in Cameroon. We seek to determine the prevalence and factors associated with PPD in women attending the Tubah District hospital, North West Region, Cameroon. We conducted a cross-sectional hospital-based study at the Tubah District Hospital. A consecutive convenience sampling technique was used to recruit participants. Our main outcome was post-partum depression which was assessed using the Edinburgh Post-partum Depression Scale. A total of 207 post-partum women took part in this study with a mean age of 27.54 ± 5.78 years. The prevalence of depression was 31.8%. Gender-based violence (OR: 4.67, P = 0.013), financial stress (OR:3.57, P = 0.002) and male baby (OR: 2.83, P < 0.001) were independent psychosocial factors associated with PPD. Independent psycho-clinical factors of post-partum depression include family history of mental health illness (OR: 4.34, P = 0.04) and previous history of depression (OR: 4.17, P = 0.02). The prevalence of post-partum depression in women attending the Tubah District Hospital, Northwest Region, Cameroon is high. The factors associated with PPD are many. Identification of risk factors, early diagnosis and proper management can prevent PPD, disabling morbidity, and suicide in mothers.

## Introduction

Bringing a child into the world is the joy of most women. However, many challenges are associated with childbirth some of which are not given appropriate attention. Amongst these is postpartum depression. Postpartum depression (PPD) is one of the mood disorders following childbirth and can develop into major depressive disorder [1,2]. Other postpartum mood disorders include baby blues and postpartum psychosis. The clinical features of PPD include anxiety, mood swings, low self-esteem, irritability, loss of appetite and lack of interest [1]. Accurate diagnosis of PPD is difficult due to similarities in its presentation with other mood disorders [3].

PPD has a very huge public health burden as shown by its high global prevalence that ranges from 4 - 63.9% [3]. In Africa previous studies have also indicated this wide prevalence range. Amongst studies conducted across Africa, a prevalence of 50.4% was recorded in the Democratic Republic of Congo [4], 34.7% in South Africa[5], 23.4 in Nigeria [6] and 6.6 % in Uganda [7]. In Cameroon, Ndjoda et al recorded 23.4% in Yaoundé [8], while Ghogomu et al, in 2016 recorded a very high prevalence of 61.8% in Limbe [9].

There are many associated factors of PPD; Gelaye et al and Stewart et al reported intimate partner violence, history of psychiatric illness, low maternal literacy levels, stress, early life abuse, anxiety, previous history of depression as predisposing factors [10,11]. In Cameroon, unemployment, unsatisfactory support of baby, marital issues, serious family problems, unplanned pregnancy, problems with baby feeding and sleep were recorded as associated factors [8,9]. Long-term negative effects can occur if PPD goes untreated. Some examples include major depression, cognitive, behavioural and physical effects which could reflect on the entire maternal family, mother and child bonding as well as child growth and development [11,12].

In sub-Saharan Africa, the focus has been more on postpartum obstetric complications and this has caused many cases of PPD to go undiagnosed and underreported, hence increasing the probability of having long term complications. Despite the magnitude of PPD, very few studies have been carried out in Cameroon. The previous studies in Cameroon were done in urban settings. No previous study has been done to describe PPD in a rural setting, so we seek to determine the prevalence and factors associated with PPD in women attending the Tubah District Hospital (TDH), North West Region, Cameroon.

## Materials and methods

### Study design, setting and participants

We conducted a cross-sectional hospital-based study from July to September 2021 at the Tubah District Hospital. All participants were women who gave birth at the hospital. The Tubah Health District is one of the 19 health districts in the Northwest Region of Cameroon. There are 13 health centres and one district hospital in the health district. The maternity of TDH carries out about 32 deliveries monthly. The activities of the maternity are run by 4 doctors, 3 midwives and 6 nurses.

### Sampling and inclusion criteria

A consecutive convenience sampling technique was used to recruit any woman who had given birth at the Tubah District Hospital. Women were recruited into the study 6 weeks after delivery at the post-natal clinic. Eligible participants who provided consent were recruited in the study. A structured questionnaire was administered to the participants by the duty midwives. The midwives had received adequate training on how to administer the questionnaire. We pre-tested our questionnaire on a group of 20 women prior to data collection. These women were excluded from the study. All poorly filled questionnaires were not included in the final analysis.

### Outcome variables

Our main outcome was post-partum depression. This was assessed using the Edinburgh Post-partum Depression Scale (EPDS). The EPDS has been used extensively to screen depression in pregnant women and post-partum women. This scale meets both DSM-5 and ICD-10 diagnostic criteria for depression[13]. EPDS is a 10-question structured screening tool that can be administered to both pregnant women and women in the post-partum period. Ideally it should be administered 4-6 weeks after delivery for the screening of post-partum depression. An EPDS score of greater than 11 is highly suggestive of a post-partum depression[13].

### Independent variables

The following variables were evaluated for association with PPD: marital status (single/divorced, married); age (years); occupation (employed and unemployed); setting (urban or rural); religion (Muslim, Christian, atheists and others); level of education; the presence of comorbidities; past history of depression and other mental health illnesses; family history of depression/mental health illnesses; alcohol consumption; cigarette smoking; HIV infection status; mode of feeding (Breast milk only, artificial/mixed feeding); sleep difficulties and pregnancy outcome (live baby, still birth).

### Sample size calculation

The sample size was obtained using the formula for estimation of a proportion since our major outcome was prevalence of post-partum depression.

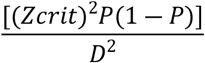

Where,

N = Number of participants

Z crit = the standard normal deviation, corresponding to a significance criterion of 0.05 (95), = 1.960

D = Amount of error we will tolerate = ± 6 %

P = Pre-study estimate of the prevalence of post-partum depression

A pre-estimate value of P=23.4 % was used. This was in accordance with a study done in an urban setting in Cameroon, where about 23.4% of study participants presented with symptoms of depression on the PHQ-9 score [8].

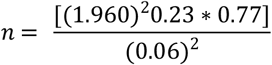

n = 188 participants.

### Statistical methods and data analysis

Data were entered into an excel spreadsheet and analysed using Stata version 14 statistical software. Results were presented as means and standard deviation (SD) for continuous variables and frequencies and percentages for categorical variables. At bivariate analysis, we used the Cochran-Mantel-Haenszel test to obtain crudes odd ratios (OR) of factors associated with PPD. Multivariate logistic regression was used to identify independent associations of PPD. This was presented as adjusted odds ratios(AOR). A p-value of <0.05 was used as cut off for statistical significance.

### Ethical considerations

We obtained ethical clearance from the Ethical Review Board of the Regional Hospital of Bamenda prior to commencement of the study. All participants provided verbal or written consent before they were recruited into the study. Assent was obtained for participants below the age of 18 years.

## Results

### Socio-demographic characteristics of participants

A total number of 207 post-partum women took part in this study with a mean age of 27.54 ± 5.78 years. Majority of the participants were employed (73.76%) and married (72.95%). Almost all participants reported the presence of sufficient social support (88.41%) or partner support (87.44%). Minority of participants reported the presence of gender-based violence (4.83%) and more than a third of them indicated some financial stress (36.10%). (Table 1).

**Table 1:** Socio-demographic characteristics of Participants

| Variables | Mean ± SD |
| --- | --- |
| Age(years) | 27.54 ± 5.78 |

|  | N | Percentage (%) |
| --- | --- | --- |
| **Employment Status** |  |  |
| Unemployed | 53 | 26.24 |
| Employed | 149 | 73.76 |
| **Education** |  |  |
| Primary | 88 | 42.51 |
| Secondary | 64 | 30.92 |
| Tertiary | 55 | 26.57 |
| **Marital Status** |  |  |
| Single/ Divorced | 56 | 27.05 |
| Married | 151 | 72.95 |
| **Gender based violence** |  |  |
| Present | 10 | 4.83 |
| Absent | 197 | 95.17 |
| **Presence of social support** |  |  |
| Present | 24 | 11.59 |
| Absent | 183 | 88.41 |
| **Supportive partner** |  |  |
| Present | 26 | 12.56 |
| Absent | 181 | 87.44 |
| **Sex of Baby** |  |  |
| Male | 116 | 56.59 |
| Female | 89 | 43.41 |
| **Financial stress** |  |  |
| Yes | 74 | 36.1 |
| No | 131 | 63.9 |

### Clinical characteristics of participants

The mean gravidity of all participants was 2.84 ± 1.80 pregnancies. The mean parity of all participants was 2.63 ± 1.53 deliveries. The mean number of ANC visits attended was 5.26±1.53 and the mean gestational age at the time of delivery was 39.25 ± 1.69. Majority of participants had a vaginal delivery (74.88%), had no post-partum complications (81.46%), were HIV negative (97.10%), had no family history of mental health conditions (92.17%). Almost all participants gave birth to live babies (94.20%) and chose exclusive breast feeding as the main mode of feeding (80.68%). (Table 2)

**Table 2:**
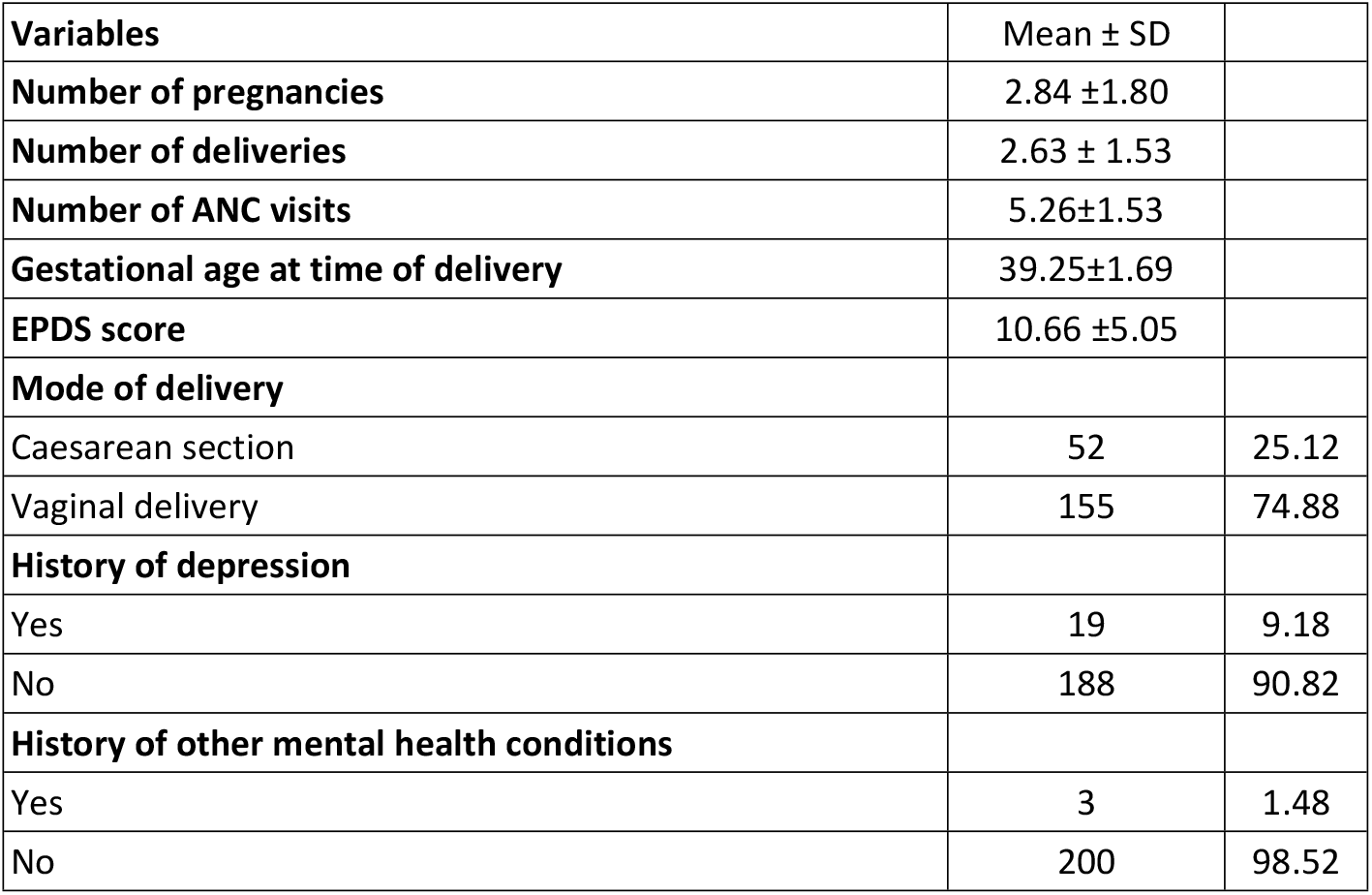
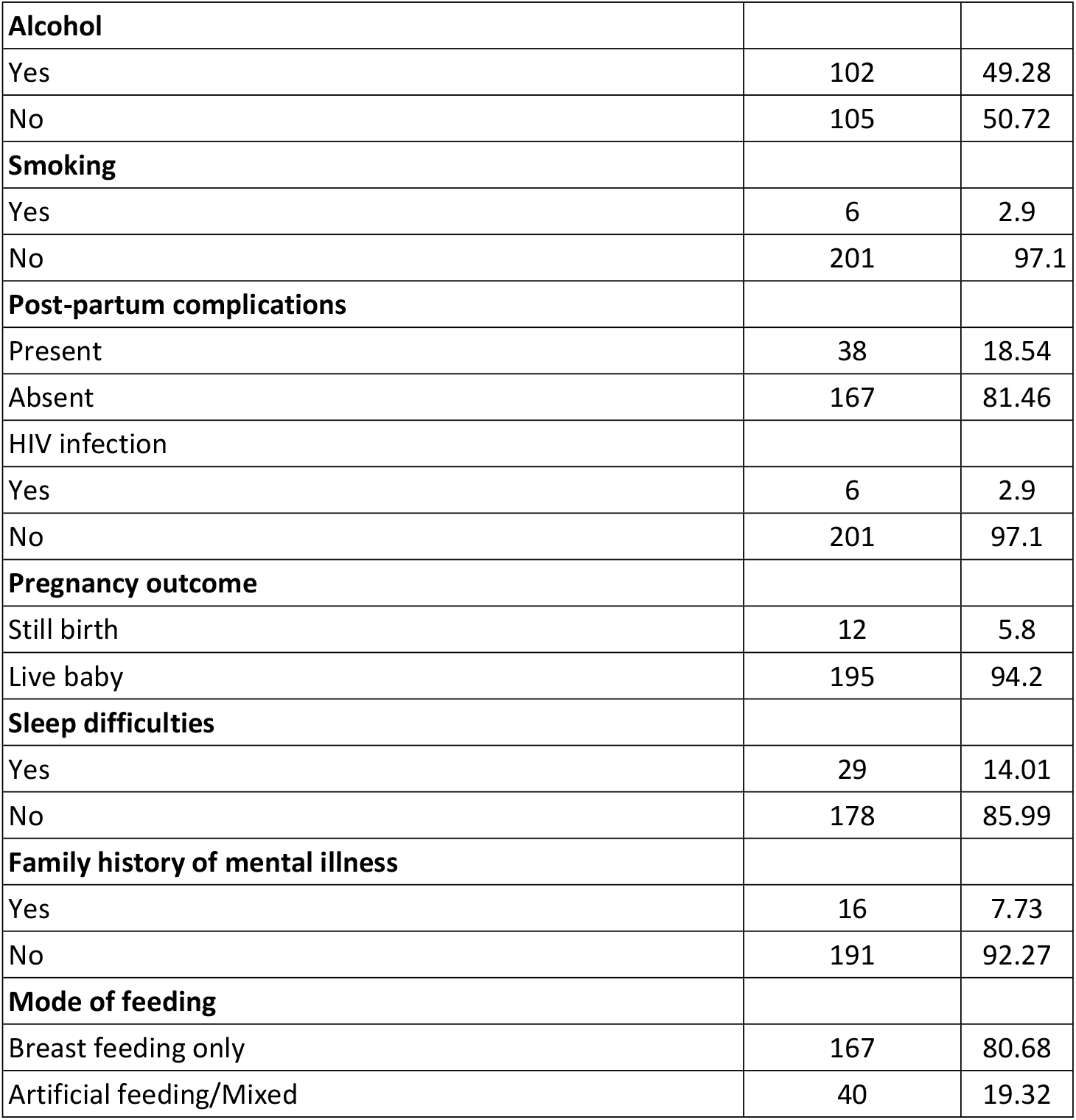
Clinical characteristics of participants

### Frequency of self-harm thoughts in participants

A total number of 123 participants reported never having any self-harm thoughts while 24 participants reported self-harm thoughts are sometimes common (Figure 1).

**Figure 1.**
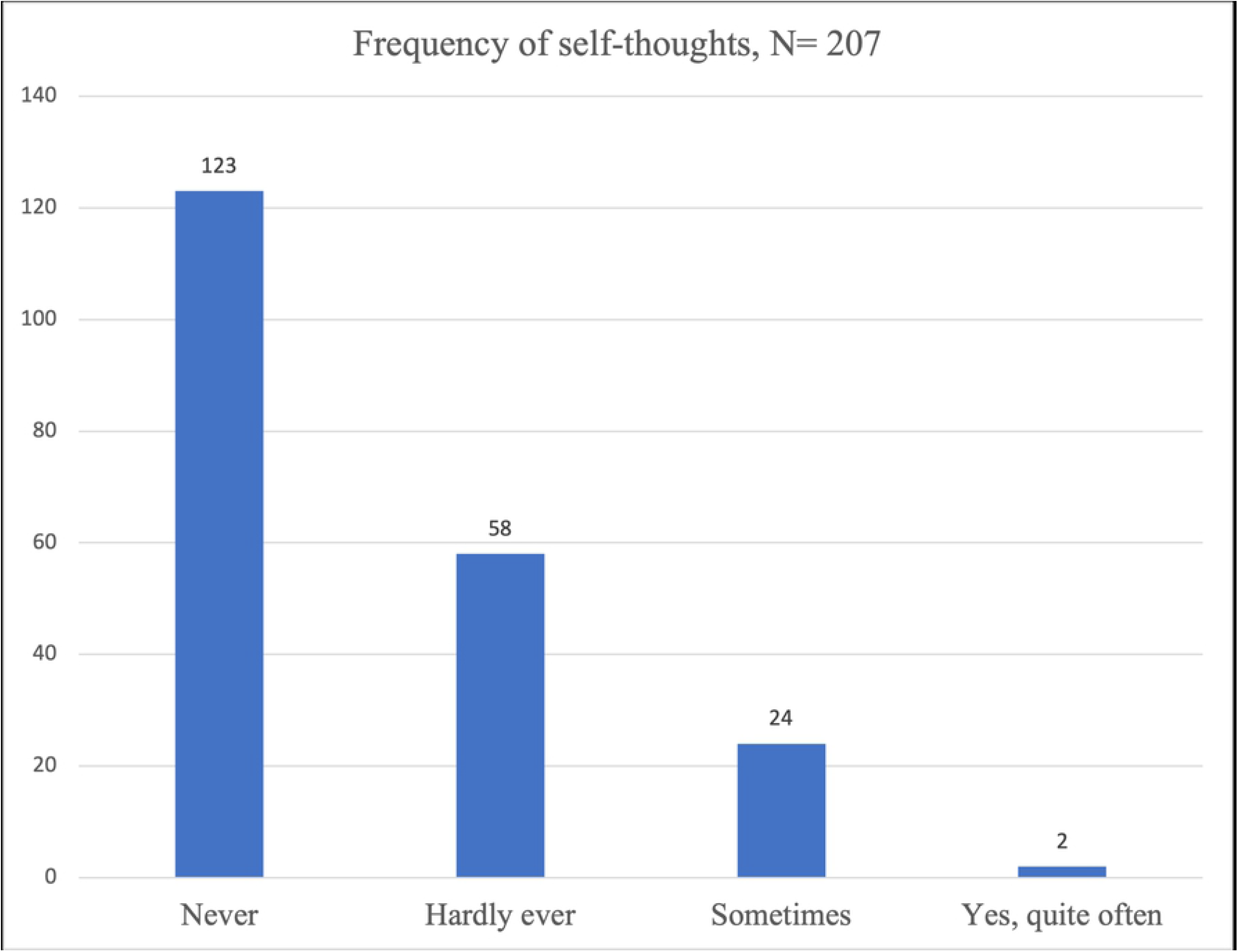

### Prevalence of depression amongst participants

The prevalence of probable depression in participants was 31.8%. (Figure 2).

**Figure 2.**
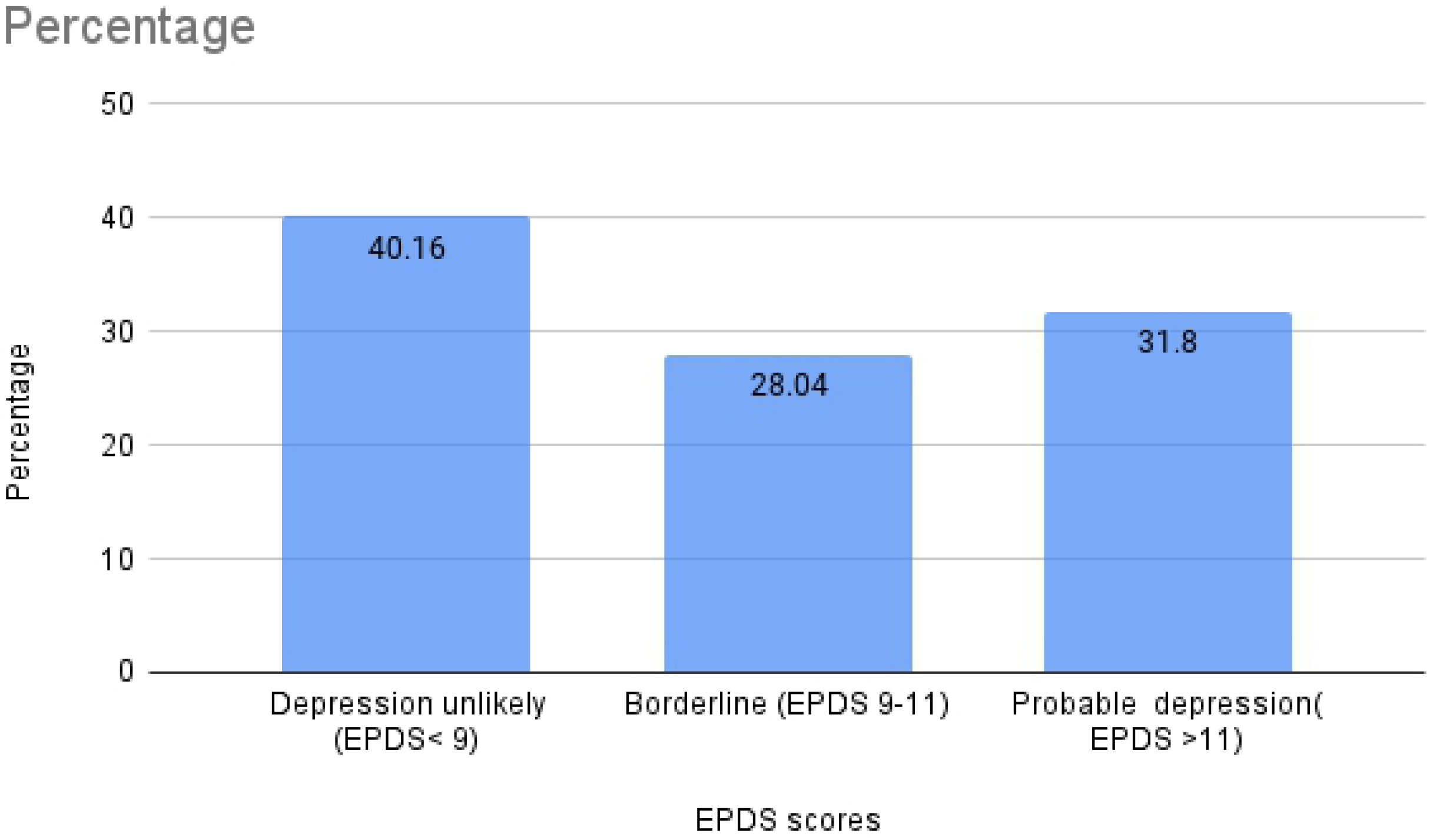

### Socio-demographic factors associated with depression

At bivariate analysis, the presence of gender-based violence (OR:5.45, P = 0.001), financial stress (OR:3.88, P < 0.001), male baby (OR: 3.09, P < 0.001) were associated with post-partum depression. Presence of a supportive partner had a protective effect against post-partum depression (OR: 0.34, P = 0.01). (Table 2). After adjusting, Gender-based violence (OR: 4.67, P = 0.013), financial stress (OR:3.57, P = 0.002) and male baby (OR: 2.83, P < 0.001). were independently associated with PPD (Table 2)

### Psycho-clinical factors associated with depression

At bivariate analysis family history of mental illnesses (OR: 3.01, P = 0.02), alcohol consumption (OR: 1.79, P = 0.04), smoking (OR: 4.48, P = 0.04) and previous history of depression (OR:4.25, P = 0.002) were associated with PPD. Also, the analysis revealed that the absence of sleep difficulties (OR:0.34, P = 0.001), and women who were already latched on (OR: 0.21, P = 0.02) were less likely to develop post-partum depression. Independent psycho-clinical factors of post-partum depression include family history of mental health illness (OR: 4.34, P = 0.04) and previous history of depression (OR: 4.17, P = 0.02). (Table 3)

**Table 3:** Psycho-clinical characteristic associated with Depression

| Variables |  | Crude ORs | P-value | Adjusted OR | P-value |
| --- | --- | --- | --- | --- | --- |
| Days of Post-partum | < = 7 | 0.95 | 0.8 | * | * |
|  | > 7 | 1 |  |  |  |
| No of ANC visits | < = 3 | 0.83 | 0.75 |  |  |
|  | >3 | 1 |  |  |  |
| Presence of Post-partum complications | Yes | 1.2 | 0.73 | * | * |
|  | No | 1 |  |  |  |
| Gravidity | <= 1 | 1.12 | 0.86 | * | * |
|  | > 1 | 1 |  |  |  |
| Parity | <= 1 | 0.97 | 0.95 | * | * |
|  | > 1 | 1 |  |  |  |
| Sleep difficulties | No | 0.34 | 0.001 | 0.2 | 0.05 |
|  | Yes | 1 |  |  |  |
| Family History of Mental disease | Yes | 3.02 | 0.02 | 4.34 | 0.04 |
|  | No | 1 |  | 1 |  |
| Alcohol consumption | Yes | 1.79 | 0.04 | 1.01 | 0.9 |
|  | No | 1 |  |  |  |
| Smoking | Yes | 4.48 | 0.04 | 1.9 | 0.1 |
|  | No | 1 |  |  |  |
| History of mental health condition | Yes | 1.92 | 0.2 | * | * |
|  | No | 1 |  |  |  |
| History of depression | Yes | 4.25 | 0.002 | 4.17 | 0.02 |
|  | No | 1 |  |  |  |
| Birth outcome | Life baby | 2.44 | 0.25 | * | * |
|  | Still birth | 1 |  |  |  |
| Milk flowing | Yes | 0.21 | 0.02 | 0.81 | 0.8 |
|  | No | 1 |  |  |  |

## Discussion

Depression is a common and debilitating complication of the postpartum period especially in Low and Middle-Income Countries (LMICS) where the burden of unmet psychological disturbances is underestimated. Abdollahi and Zarghami noted that women who experienced an episode of PPD suffered adverse outcomes years later; they were more likely to be depressed, suffer from chronic diseases, and scored higher on the general health questionnaire [14]. Similarly, the outcome of PPD in children has been well documented; they experience higher rates of behavioural problems, delayed cognitive and language development, poor academic grades, and depression later in life [15]. Thus, given the adverse long-term outcomes of PPD on both mother and child, it raises a major concern and calls for therapeutic action. The aim of this study was to determine the prevalence and factors associated with PPD among women attending post-natal clinic at the Tubah district hospital, North West Cameroon.

The prevalence of PPD in this study was 31.8%. This falls within the range of 4.0% - 63.9% reported by Arifin SRM et al [16] in postpartum mothers globally especially in Sub-Saharan Africa and the 6.9% to 50.3% obtained by Atuhaire et al [17] among mothers in Africa only. This rate is comparable 31.7% obtained by Hung et al in South Africa [18] and 33% by Chibanda et al in Zimbabwe [19]. However, this prevalence is higher than that reported by Nakku et al in Uganda (6.1%) [7], Anokye et al in Ghana (7%) [20], Adewuya et al in Nigeria (13.1%) [6], and Toru et al in Ethiopia (22.9%) [21]. The different tools used (SRQ-25, PH-9, ZSDS, and PHQ-9) could explain this variation. These tools are less specific and their use could extend to 12 months post-partum unlike the recommended 6 to 12 weeks for the EPDS[15].

On the contrary, the prevalence of PPD in our study was lower than that reported by Ghogomu et al, in Limbe Cameroon (61.8%) [9]. This disparity could be explained by the fact that his study was community-based and assessed mothers up to 12 months postpartum there by exposing the mothers to other risk factors of depression other than childbirth. Ideally, mothers should be screened for PPD one month postpartum or at 6 weeks [22]. It is worthy of note that this study was done in the North West region of Cameroon which has suffered socio-political instability for over 5 years now. This has hindered access to healthcare by many and therefore the prevalence obtained in our study might be an underestimate of the actual situation. Hence, there is a need for more community-based studies in this era of socio-political instability to have a better appraisal of this major postpartum complication. In addition to socio-political instability, the ongoing Covid-19 pandemic might have also contributed to our higher rates of PPD, an observation shared elsewhere by Liang et al [23].

Financial stress was associated with PPD in our study. This is similar to findings reported by Djoda et al [8] and Ghogomu et al [9]. Participants who experienced gender-based violence were more likely to be depressed. This is congruent with findings reported in the literature [8– 11]. The occurrence of PPD was high in those with a family history of mental illness. This is supported by findings from Stewart et al [11] and Neslihan et al [24]. Similarly, a history of maternal depression was associated with PPD in our study as was the case in the review by Neslihan et al [24].

Furthermore, the male gender was associated with PPD in our study. This is contrary to what Amr et al had in Saudi Arabia [25] as they found the birth of a female baby to be associated with PPD. This could be explained by the cultural differences in the study areas; Saudi Arabia is a predominantly muslim community where the birth of a male child is celebrated more than a female child. More so it is a patrilineal society where the issue of an heir is considered of great importance.

It is important to consider these results within the context of the study’s limitations: The fact that it was a single centre hospital-based study may have influenced the highly selected group of participants. It may therefore be difficult to generalise the findings to the entire Cameroonian population. This could be improved by carrying out community-based studies and multicentre hospital-based studies. Also, the self-reported nature of the questionnaire and some elements of the EPDS scale leave room for recall bias and socially desirable responses. This score is a screening tool that validates the need for a psychiatrist to confirm the diagnosis which was not done in our study. More so, the list of factors potentially associated with PPD as evaluated in this study is by no means exhaustive.

Despite these limitations, to the best of our literature search, this is the first study that provides evidence-based data on the prevalence and factors associated with PPD in a rural area in Cameroon. Whilst we envisage the scalability of this project, the observed high rates of PPD herein highlight the necessity for tailored pre- and post-partum counselling and monitoring from qualified personnel. However, the responsibility of ensuring a smooth post-partum transition does not entirely rest on healthcare professionals as evidence suggests good family/partner support is invaluable. This therefore calls for collective efforts if we are to curb the deleterious long-term effects of post-natal depression on the mother which will intend enhance proper growth and development of the newborn.

## Strengths and limitations

The prevalence of PPD in urban Cameroon ranges from 23.4 to 61.8% in 2 previous studies. Psychosocial factors including unemployment, unsatisfactory support of baby, marital issues amongst others are associated with PPD. This study to the best of our knowledge this is the first study to determine the prevalence of PPD in a rural setting in Cameroon (31.8%) thus providing relevant evidence-based data which could help inform policy.

## Conclusion

The prevalence of PPD was found to be 31.8% in a rural primary care centre population of childbearing women in Cameroon. The psycho-clinical factors associated with PPD were a family history of mental illness and a history of depression, while the social factors associated with PPD were gender-based violence, financial stress and having a male baby. These findings will be fundamental in designing antenatal and postnatal screening tools that will be used by health care providers to screen for these psycho-clinical and social factors to prevent PPD, disabling morbidity, and suicide in mothers.

## Data Availability

All relevant data are within the manuscript and its Supporting Information files.

## Competing interests

The authors declare no competing interest

## Authors’ contributions

SNN and TND conceptualised the project and designed the protocol and questionnaires and participated in the writing of the article. NCN ran the analysis of the collected data and wrote certain sections of the article. CA, ET, MB, JON, LAST and CSB contributed to the write-up of the different sections of the article. All authors read and approved the final copy of the manuscript

## Acknowledgements

Our sincere gratitude goes to all the staff of the TDH Maternity and the midwives who helped immensely with the data collection of this study. In addition to previous psychosocial factors described in the literature, our study found out that women who had a male baby had higher odds of having PPD

## Notes

### Competing Interest Statement

The authors have declared no competing interest.

### Funding Statement

The author(s) received no specific funding for this work

